# Deep Learning based retinal layer segmentation in optical coherence tomography scans of patients with inherited retinal diseases

**DOI:** 10.1101/2023.09.26.23295852

**Authors:** Franziska Eckardt, Robin Mittas, Nastassya Horlava, Johannes Schiefelbein, Ben Asani, Stylianos Michalakis, Maximilian Gerhardt, Claudia Priglinger, Daniel Keeser, Nikolaos Koutsouleris, Siegfried Priglinger, Fabian Theis, Tingying Peng, Benedikt Schworm

**Affiliations:** Department of Ophthalmology, LMU University Hospital Munich, LMU Munich, Mathildenstrasse 8, 80336 Muenchen, Germany; Helmholtz Munich, Ingolstaedter Landstrasse 1, 85764 Neuherberg; Department of Psychiatry and Psychotherapy, LMU University Hospital Munich, LMU Munich, Nußbaumstrasse 7, 80336 Muenchen, Germany

## Abstract

**Background:** On optical coherence tomography (OCT) scans of patients with inherited retinal diseases (IRDs), the outer nuclear layer (ONL) thickness measurement has been well established as a surrogate marker for photoreceptor preservation. Current automatic segmentation tools fail in OCT segmentation in IRDs, and manual segmentation is time consuming.

**Methods and Material:** Patients with IRD and the availability of an OCT scan were screened for the present study. Additionally, OCT scans of patients without retinal disease were included, to provide training data for the artificial intelligence (AI). We trained a U-net based model on healthy patients and applied a domain adaption technique to IRD patients’ scans.

**Results:** We established an AI-based image segmentation algorithm that reliably segments the ONL in OCT scans of IRD patients. In a test dataset, the dice-score of the algorithm was *98.7%*. Furthermore, we generated thickness maps of the full retinal thickness and the ONL layer for each patient.

**Conclusion:** Accurate segmentation of anatomical layers on OCT scans plays a crucial role for predictive models linking retinal structure to visual function. The here-presented OCT image segmentation algorithm could provide the basis for further studies on IRDs.

## Introduction

Inherited retinal diseases (IRDs) are a diverse group of genetic disorders that lead to progressive degeneration of the retina. These diseases encompass a wide range of conditions such as Retinitis Pigmentosa, Leber’s Congenital Amaurosis, Stargardt disease, and many others. IRDs often lead to significant vision impairment and potentially blindness. In Europe, IRDs affect about 1:3000 individuals. Optical Coherence Tomography (OCT), as a noninvasive imaging test, has become an indispensable tool for diagnosing and monitoring the progression of retinal conditions for IRDs [1,2]. OCT imaging allows an analysis and measurement of the retina’s distinct layers, which can be crucial for confirming diagnosis, guiding treatment plans, and assessing response to therapies in the management of IRDs.

With the advent of artificial intelligence (AI) and machine learning, there have been several studies aiming to help disease diagnosis based on OCT. However, compared to more common ophthalmological diseases like diabetic retinopathy (DR) [3], age related macular degeneration (AMD) [4] and glaucoma [5], AI studies specialized on IRDs are still scarce. In this work, we initiated a study for a deep learning based retinal layer segmentation for IRD patients with a special focus on the outer nuclear layer (ONL). In IRDs, ONL thickness measurement has been well established as a surrogate marker for photoreceptor preservation. The goal of this work was to establish an AI-based image segmentation algorithm that reliably segments the ONL in OCT scans and provides a full retina and ONL thickness map of IRD patients.

## Methods and Material

### Participants and imaging

The clinical research database of the Department of Ophthalmology was screened for patients with a confirmed IRD diagnosis and available OCT scans. Normal data was collected from healthy participants without retinal disease. From every individual both eyes were included in this study. Spectral domain optical coherence tomography (OCT) and near infrared (NIR) confocal scanning-laser ophthalmoscopy was performed using the Spectralis HRA+OCT platform (Heidelberg Engineering GmbH, Heidelberg, Germany). Each OCT scan consisted of either 49 or 97 slides (one slide refers to one 2D image (B-scan) with a resolution of 496x512 pixels) and covers 20x20 degrees of the posterior pole centered to the fovea centralis. OCT scans were only included with a quality index of 20 or better and no blinking artifacts. OCT scans were exported as .dcm files from the manufacturer’s software Heidelberg Eye Explorer (Version 1.10.4.0, Heidelberg Engineering GmbH, Heidelberg, Germany). The local Ethics Committee of the Medical Faculty gave ethical approval for this work (identifier 23-0392). The study adhered to the tenets of the Declaration of Helsinki.

### Manually annotated OCT-segmentation for training datasets

The annotations for the healthy retinal dataset were obtained from the software "OCTExplorer", which was developed by the IOWA University [6–8] and is free-to-use by academics. We used this software as its segmentation results on healthy retinal OCTs are reliable and only a small proportion of slides needed to be re-annotated by an expert. Furthermore, it is also possible to access the segmented files from the software to further process and transform the segmentation by a small helper function.

However, in IRD patients, the existing automatic segmentation tools, including the above-mentioned "OCTExplorer", fail to reliably segment these scans, due to structural alterations caused by the degeneration of retinal layers. Examples of OCT Explorer segmentations are shown in *Figure 1* and *2*.

**Figure 1:**
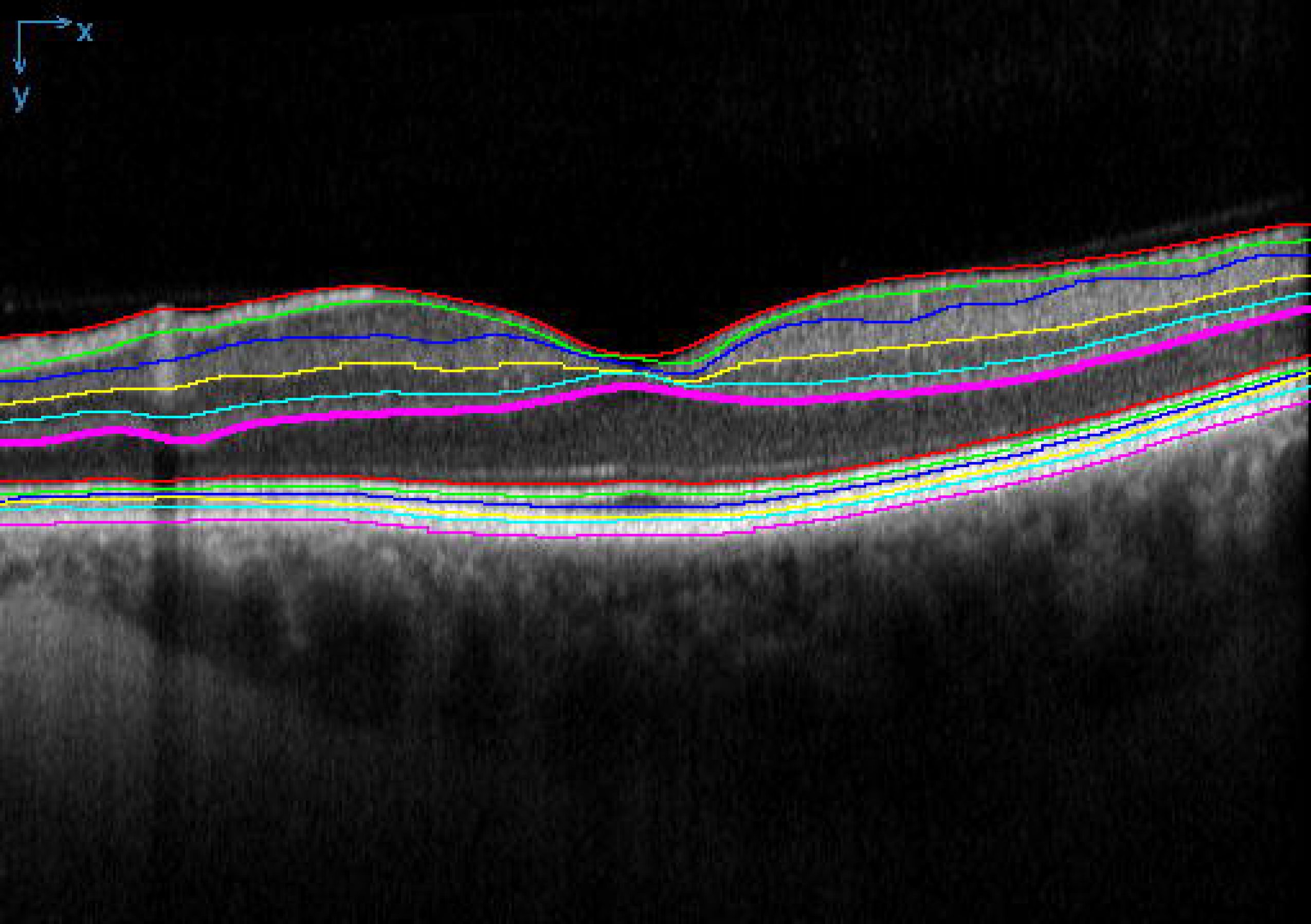
OCTExplorer segmentation screenshot for a healthy patient. Bold: segmented outer plexiform layer (OPL)-Henle fiber layer. OCT = optical coherence tomography; IRD = inherited retinal disease

**Figure 2:**
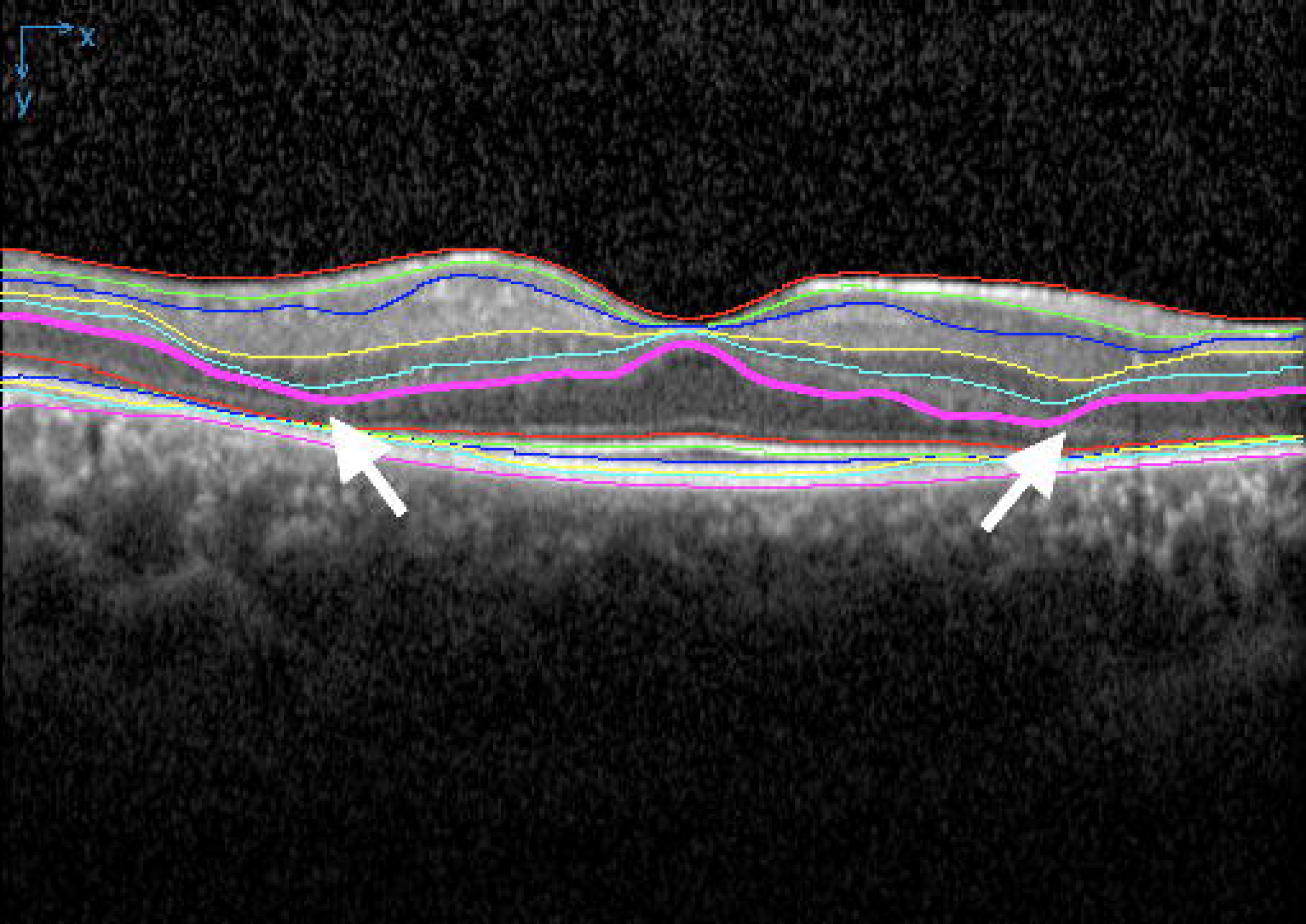
OCTExplorer segmentation screenshot for an IRD patient. Bold: segmented outer plexiform layer (OPL)-Henle fiber layer. Note that the bold pink line for the segmentation of the OPL-Henle fiber layer boundary crosses over to the actual inner nuclear layer (white arrows). OCT = optical coherence tomography; IRD = inherited retinal disease

Consequently, we had to rely on expert annotations as our ground-truth for IRD patients, which is a laborious and time-consuming process. OCT scans were graded from two expert ophthalmologists. For IRDs, the segmentation of the outer retinal layers, especially the outer nuclear layer (ONL), is essential to monitor structural degeneration over time. Thus, in our study, the focus was laid on a precise segmentation of the ONL. Within this work we will refer to the convex thickening of the ONL within the foveal pit as “ONL-hill” on central OCT slides as depicted in *Figure 3*.

**Figure 3:**
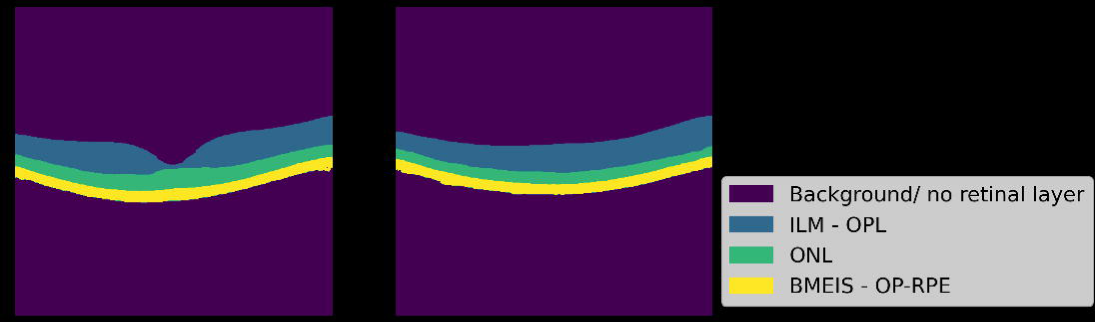
(a) Exemplary slide in the middle of an OCT scan in a healthy patient with the ONL-’’hill’’ (b) Exemplary first slide of an OCT scan without ONL-’’hill’’. The same color codes for retinal layers will be used within this work. OCT = optical coherence tomography; ONL = outer nuclear layer

### Development of the deep learning-based segmentation algorithm and statistical analysis

To our best knowledge all existing deep learning-based approaches to segment retinal layers of OCT scans are based on a U-net architecture, which was also the choice for our model. Here, we rely on the Python module *Tensorflow* for our model definition and training. The versions of all packages we utilized can be found on <u>GitHub</u> in the requirements file. We ran the code within the de.NBI-cloud (Deutsches Netzwerk für Bioinformatik-Infrastruktur) to which we uploaded anonymized input images. The “de.NBI Cloud Berlin - Production” has provided us with sufficient compute instances for our project.

Since our OCT scan has an an-isotropic resolution, we used 2D convolutional kernels rather than 3D kernels and processed the scan in a slide-by-slide fashion. Specifically, the backbone network consists of five symmetrical pairs of encoding and decoding blocks, which perform contraction and expansion operations, respectively, during the data feedforward process. Each encoding/decoding block is composed of two convolutional layers with (3 x 3) convolution kernels, followed by a batch normalization layer, an exponential linear unit (ELU) function and a (2 x 2) max-pooling operation with stride 2. The first encoding block results in 16 feature channels, and every subsequent encoding block doubles the number of channels. Each decoding block is symmetric with respect to the encoding block at the corresponding level, except that the max pooling layer is replaced with transposed convolutional layers to upsample the feature map. In the last decoding block, we added a (1 x 1) convolutional layer followed by Softmax activation function to map the 16 feature channels to a four-class probability map that softly assigns each pixel to the background or one of the three foreground retinal layer classes, namely the layers given in *Table 1*. The model architecture is very similar to [9], which has shown to work for the task of brain tissue segmentation of different modalities/species. Instead of using a bottleneck dimension of 512, we have used a dimension of 1024. Further we did not include the nonlocal attention block in our model as this additional layer did not increase the model’s performance in our task. Lastly, as data augmentation during training we have added random gaussian noise to the input 2D slides.

**Table 1:** Proportion of pixels of retinal layers of the healthy vs. IRD patient’s datasets. As compared to healthy retinas, the ONL in IRD patients is considerably lower (4.0% vs. 0.5%). Internal Limiting Membrane (ILM); Outer Plexiform Layer (OPL); Henle Fibre layer (HFL); Outer Nuclear Layer (ONL); Boundary of myoid and ellipsoid of inner segments (BMEIS); Outer boundary of retinal pigment epithelium (OB-RPE).

| Retinal layer | Healthy OCTs | IRD OCTs |
| --- | --- | --- |
| Background | 83.8% | 87.3% |
| ILM-OPL-HFL | 8.8% | 10.3% |
| ONL | 4% | 0.5% |
| BMEIS-OB-RPE | 3.3% | 1.8% |

Our objective loss function is a combination of the cross-entropy loss and the multiclass dice coefficient loss. For IRD patients, the ONL layer is much thinner as compared to healthy subjects. Therefore, we introduced a different weight for different layers within the loss function, penalizing the ONL layer 4 times higher than other layers for IRD patients, as our primary goal is to accurately segment the ONL. Here, we used one hot encodings of shape (4, 496, 512). The very last layer uses the Softmax activation function, returning the probability for each pixel belonging to one of the four classes.

To compensate for the limited manual annotations available for IRD patients, we utilized the wealth of automatic annotations from normal OCT scans generated by "OCTExplorer". By employing domain transfer and transfer learning techniques, we bridged the gap between IRD and standard OCT scans. This domain transfer network was originally developed by Ziqi Yu et al. [9] for Brain MRI segmentation, as described in detail in [9]. Essentially, we repurposed the weights of the model trained on healthy patients - we have frozen the weights of all trainable layers, except for all batch normalization (Batchnorm) layers, resulting in adaptive Batchnorm modules. By adapting this approach, we successfully achieved efficient learning using a small number of annotations and overcame the domain shifts between IRD and normal OCT scans. However, we also compared models which did not freeze the weights of all layers.

## Results

### Dataset characteristics

OCT scans from 12 healthy control individuals without retinal disease were collected, two OCT scans each, one for the left and right eye, respectively. For the healthy control training dataset, 18 volume scans consisting of 49 B-scans each from 12 patients met the quality criteria and were included. For the IRD training dataset, 16 patients were selected from the database. Because some OCT volume scans failed to meet the quality criteria, we included 25 volume scans in total, each consisting of either 49 or 97 B-scans.

Due to the degenerative nature of IRDs, tissue loss becomes also evident by the unbalanced pixel-proportion of the total scan area for different retinal layers, as shown in *Table 1*. Particularly, the ONL is considerably reduced compared to healthy controls (from 4.0% to only 0.5%).

For all results, we trained the U-net based model for 30 epochs with early stopping. For all models trained on healthy or diseased patients we used the same hold out validation and test sets. Furthermore, we split training, validation, and test sets by patient IDs, such that there was no overlap between training and validation sets. Lastly, we used 2D-slides as input to the model. The test and validation set consisted for both, healthy and IRD datasets, each of scans of *2* patients’ volume scans, mostly including a scan of the left and right eye respectively. The codes and our trained models can be found on GitHub (https://github.com/peng-lab/retina-segmentation).

### Results on healthy patients

First, we trained a model on the full training dataset, consisting of OCT scans of healthy individuals. Here, our model resulted in a Dice Coefficient of *99,4%* for a hold-out test set. Exemplary segmentation results are illustrated in *Figure 4* and *Figure 5.* It is noteworthy that we just included OCT scans which were labeled and verified by our expert annotators.

**Figure 4:**
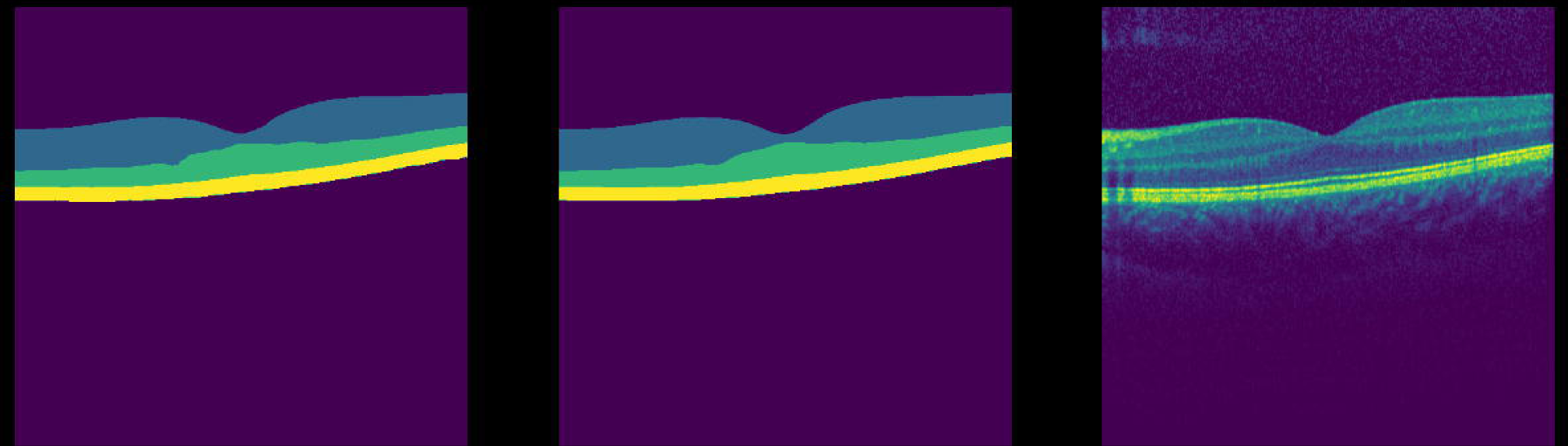
Slide containing the ‘’ONL’’ hill (a) Prediction of the model. (b) Ground truth. (c) Input slide. ONL = outer nuclear layer

**Figure 5:**
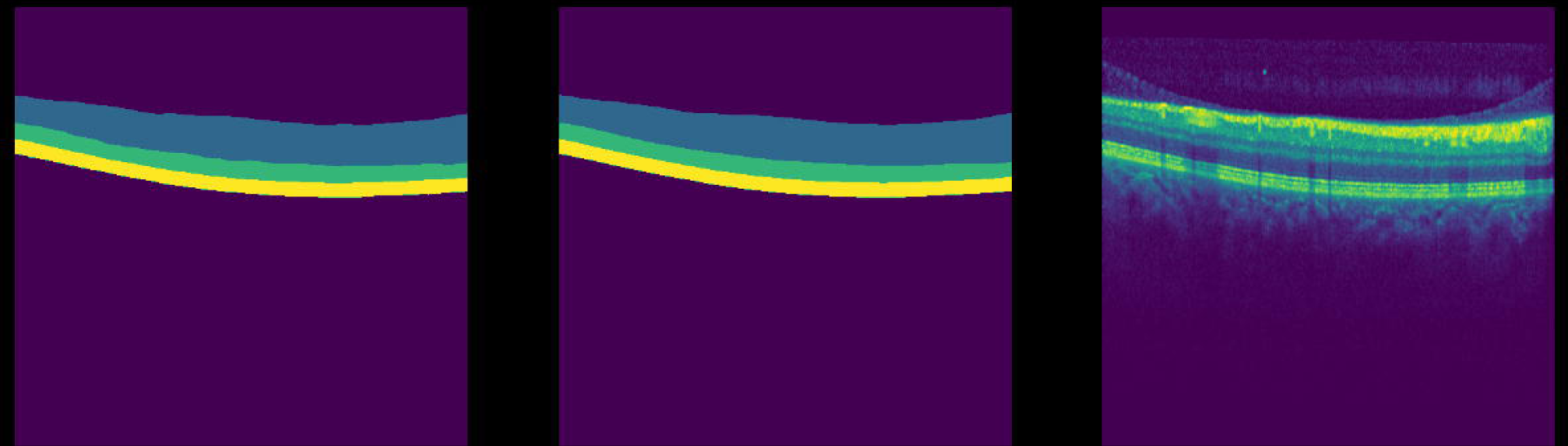
Regular slide. (a) Prediction of the model. (b) Ground truth. (c) Input slide.

We also trained models containing the nonlocal block, as described in [9], but it does not outperform the model not including this additional layer quantitatively. Further, it led to occasionally isolated misclassified pixels. Eventually we decided to not include the nonlocal block.

### Results on IRD patients

To segment retinal layers of IRD patients we tried a few different approaches: *Model 1* trained with the same loss function as for healthy patients with randomly initialized weights, (2) *Model 2* trained by upweighting the ONL layer in the Cross-Entropy loss with randomly initialized weights, (3) *Model 3* directly applying the learned model of healthy subjects on IRD patients and (4) *Model 4* using domain adaption (DA) by freezing all weights apart of the Batch-Normalization layers of the model trained on healthy subjects (*Adaptive Batchnorm*) and (5) *Model 5* trained with transfer learning by reusing the weights of the model trained on healthy individuals for initialization, however, without freezing the weights of all layers. Notably, models reusing weights from the previous model were fitted faster. The corresponding Dice coefficients can be found in Table 2.

**Table 2:** Dice-Coefficient results for segmentation on the IRD dataset. IRD = inherited retinal disease

| **Model** | **Description** | **Dice-coefficient** | **Dice-coefficient ONL** |
| --- | --- | --- | --- |
| *Model 1* | None-weighted ONL loss | 98.5% | 44.2% |
| *Model 2* | Weighted ONL loss | 98.5% | 67.4% |
| *Model 3* | Direct application of healthy model | 92.1% | 31.7% |
| *Model 4* | DA by freezing trainable weights | 98.8% | 70.2% |
| *Model 5* | Transfer learning without freezing weights | 98.8% | 70.1% |

*Model 1* failed to correctly segment the middle slides with the convex curvature of the ONL within the foveal pit. Therefore, we have introduced a weighted loss for the ONL layer for all remaining models. We have found that a weight of 4 provides the best results to accurately segment the ONL. This determination was made through hyperparameter optimization. From *Model 2* we concluded that upweighting the ONL layer in the Cross Entropy loss helped to better capture and segment this layer and it further increased the overall Dice-coefficient. Without using the weighted loss, the model oftentimes did not capture the ONL layer at all for IRD patients, as this layer is nearly non-existent in many slides. Therefore, the model ended up not assigning any probability to the ONL layer.

Furthermore, we could conclude that we needed two distinct models to segment OCT scans of IRD and healthy individuals. Directly applying the model trained on healthy individuals, *i.e. Model 3,* lead to poor performance compared to other models. The models that exhibited the highest performance were those that upweighted the ONL within the Cross-Entropy loss. However, by using the already learned features from the model trained on healthy control individuals, the model could increase its performance. *Models 4* and *5* exhibited the highest Dice-scores while they were simultaneously trained more quickly in comparison to *Models 1* and *2*. The optimal performance was attained by the Domain adaption model, which uses the *Adaptive Batchnorm* approach. Consequently, we opted to use this model for visualization purposes. *Figures 6, 7* and *8* show segmentation results for input slides from our test set. Notably, if the quality of the scan is low, the model occasionally ended up misclassifying a few random pixels as depicted in *Figure 9*.

**Figure 6:**
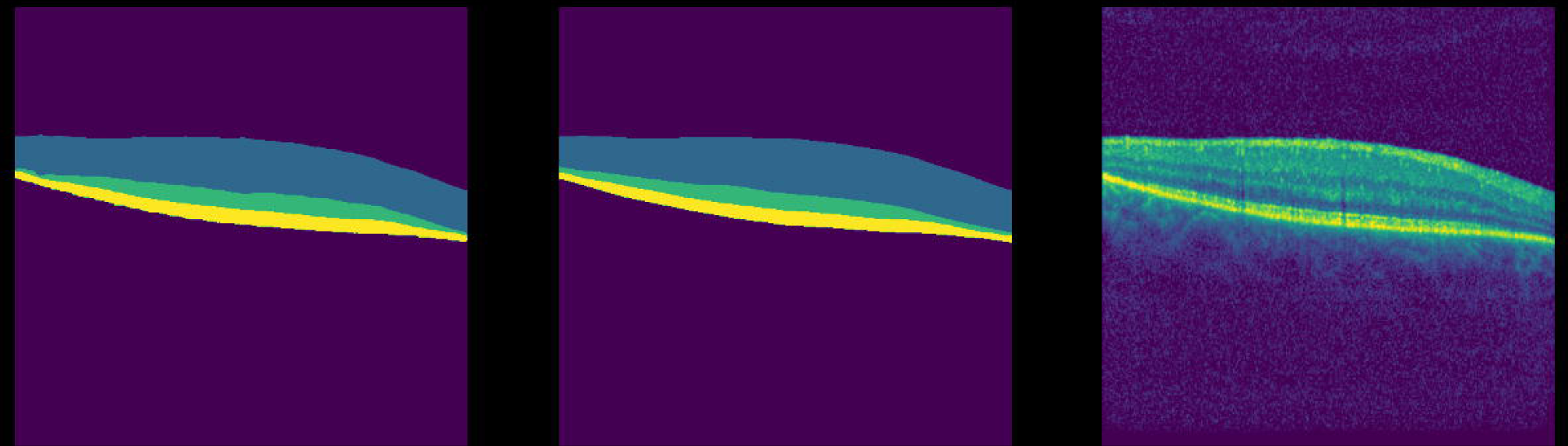
Trained model on the IRD dataset. (a) Prediction of the model. (b) Ground truth. (c) Input slide. IRD = inherited retinal disease

**Figure 7:**
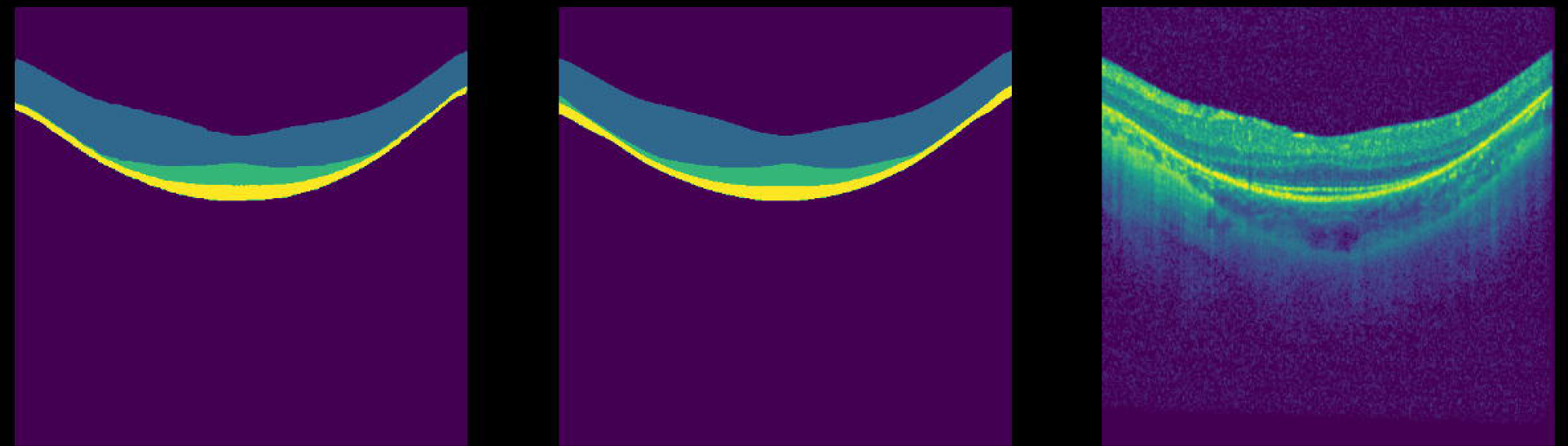
Trained model on the IRD dataset where ONL hill is visible. (a) Prediction of the model. (b) Ground truth. (c) Input slide. IRD = inherited retinal disease; ONL = outer nuclear layer

**Figure 8:**
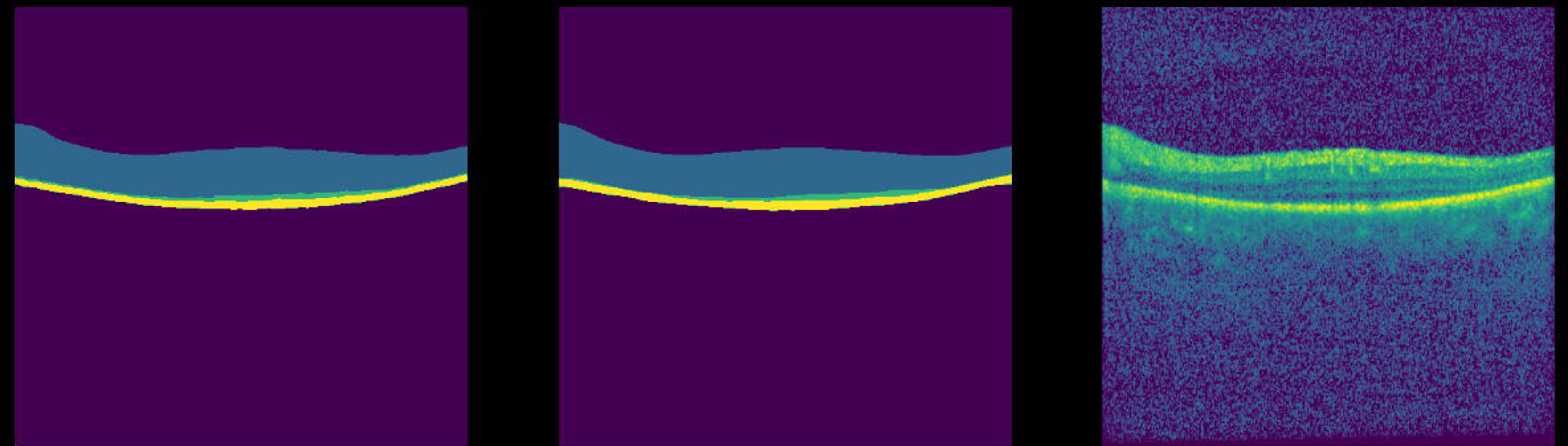
Trained model on a IRD dataset where ONL layer is almost vanished. (a) Prediction of the model. (b) Ground truth. (c) Input slide. IRD = inherited retinal disease; ONL = outer nuclear layer

**Figure 9:**
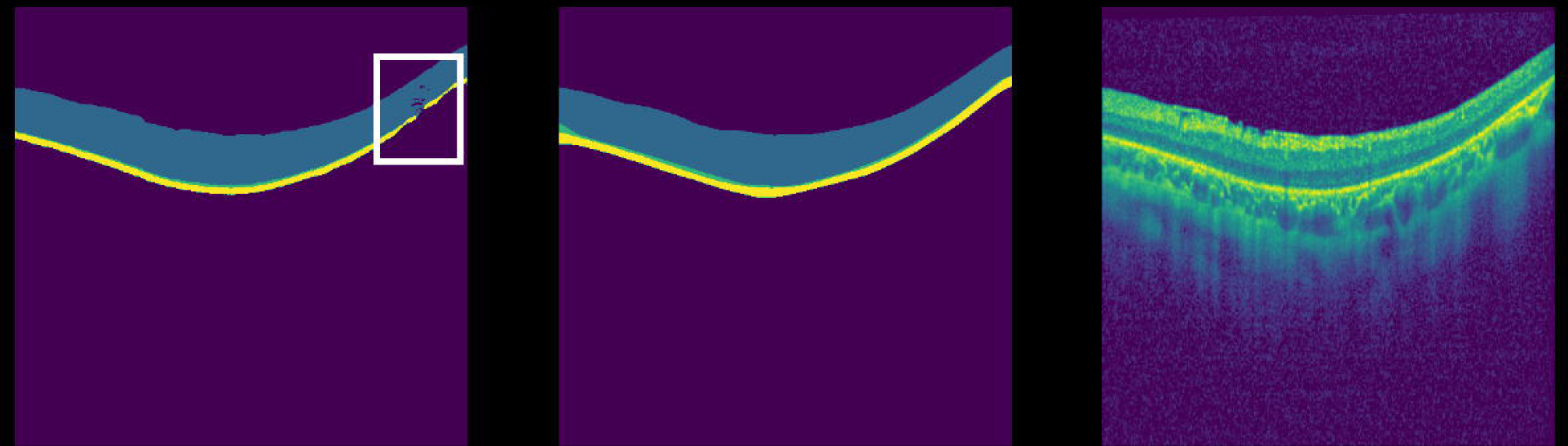
Trained model on a IRD dataset: Model has problems when quality is too low. White frame indicates some wrongly segmented pixels. (a) Prediction of the model. (b) Ground truth. (c) Input slide. IRD = inherited retinal disease

We also worked on a grid-based visual analysis using the widely used ETDRS (Early Treatment Diabetic Retinopathy Study) regions as depicted in *Figure 10*. Based on the segmentation results, we calculated thickness maps of all retinal layers or of individual retinal layers. Exemplary thickness maps including the ETDRS regions/ grid are depicted in *Figures 11* and *12*.

**Figure 10:**
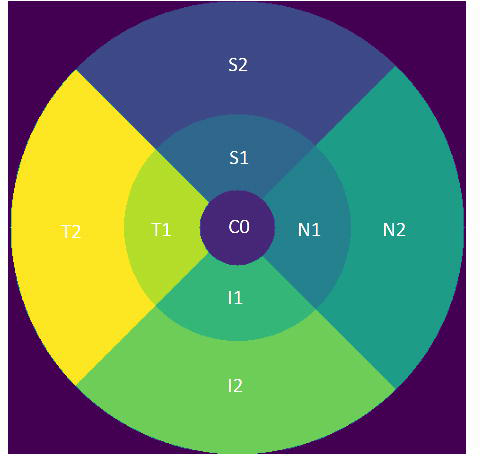
ETDRS regions.

**Figure 11:**
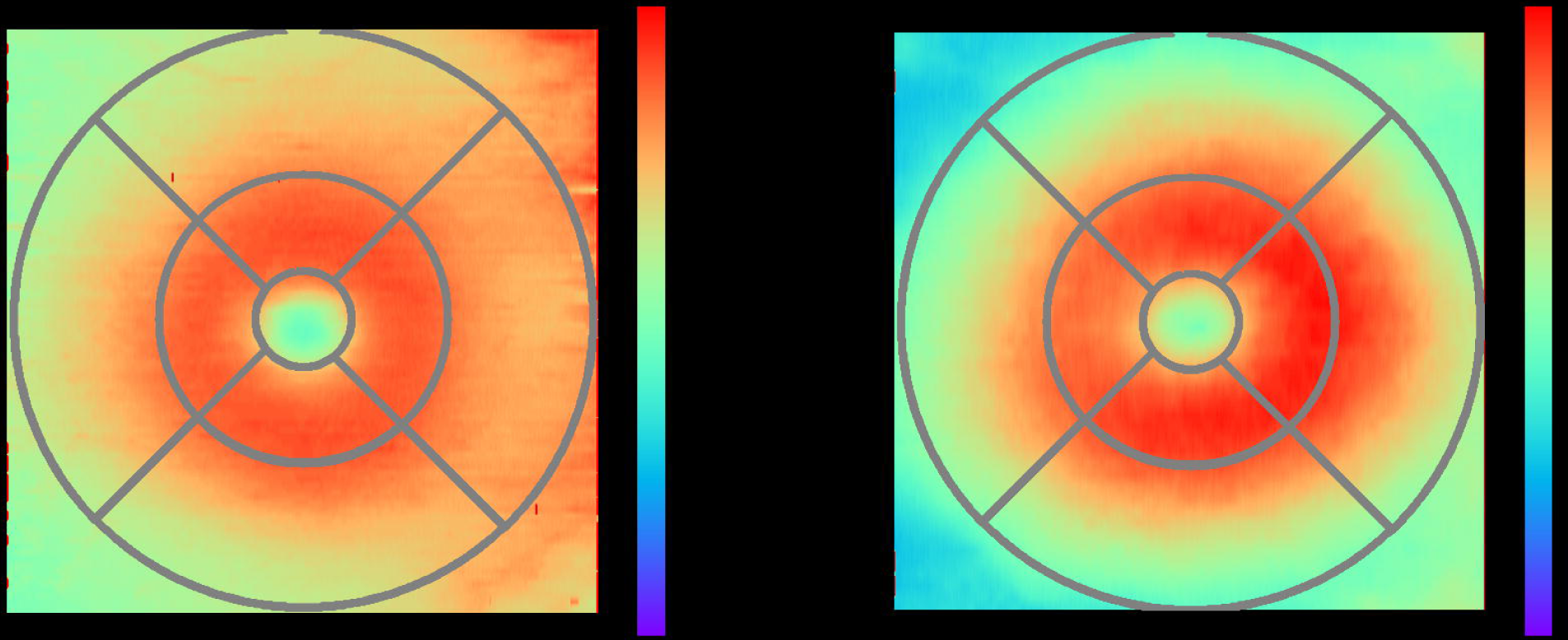
Thickness maps based on predicted segmentation of all retinal layers. (a) Thickness map of a healthy individual. (b) Thickness map of an IRD patient. Interestingly, the diseased individual has a similar full-retinal thickness. Healthy: C0 Average thickness: 274µm; S2 Average thickness 301µm; S1 Average thickness 354µm; N1 Average thickness: 352µm; N2 Average thickness: 325µm; I1 Average thickness: 352µm; I2 Average thickness: 302µm; T1 Average thickness: 348µm; T2 Average thickness: 294µm Diseased: C0 Average thickness: 281µm; S2 Average thickness 270µm; S1 Average thickness 359µm; N1 Average thickness: 369µm; N2 Average thickness: 310µm; I1 Average thickness: 366µm; I2 Average thickness: 279µm; T1 Average thickness: 343µm; T2 Average thickness: 278 IRD = inherited retinal disease

**Figure 12:**
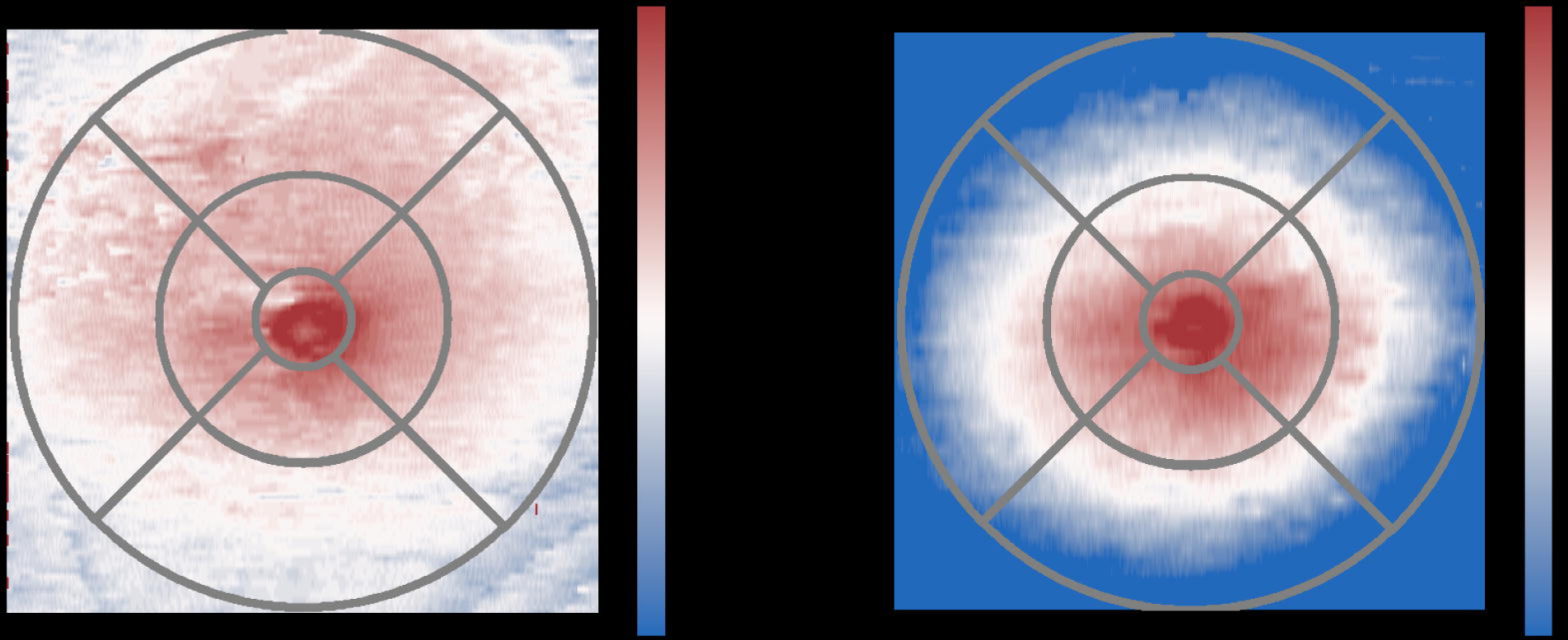
Thickness maps based on predicted segmentation of ONL layer. (a) Thickness map of a healthy individual. (b) Thickness map of an IRD patient. Evidently, the IRD patient has a thinner ONL thickness, especially at the non-central regions. Healthy: C0 Average thickness: 109µm; S2 Average thickness 80µm; S1 Average thickness 88µm; N1 Average thickness: 94µm; N2 Average thickness: 75µm; I1 Average thickness: 88µm; I2 Average thickness: 68µm; T1 Average thickness: 89µm; T2 Average thickness: 76 µm Diseased: C0 Average thickness: 115µm; S2 Average thickness 37µm; S1 Average thickness 81µm; N1 Average thickness: 94µm; N2 Average thickness: 48µm; I1 Average thickness: 92µm; I2 Average thickness: 42µm; T1 Average thickness: 89µm; T2 Average thickness: 51 µmro IRD = inherited retinal disease

## Discussion

The aim of the present study was to build an AI-based tool for the segmentation of OCTs in IRD patients, where currently available algorithms fail. Additionally, we developed a full-retina and ONL thickness map, which provides a fast overview of the important retinal layers in IRD.

With the advent of AI and machine learning, several studies aiming to enhance diagnosis, prognosis, and treatment options for ophthalmological diseases have been published [10–14]. Evaluation of retinal imaging is essential in the diagnosis of these diseases. It is a time-consuming process that requires specialists and may show variable interpretation depending on the examiner [14]. OCT allows non-invasive structural retinal imaging and generates important information about disease development and treatment response. AI-based accurate OCT analysis could be a huge advantage and potentially prevent individual interpretation errors depending on the investigator.

However, compared to more common ophthalmological diseases, AI studies specialized on IRDs are still scarce. A systemic PubMed search found only two existing works for deep learning-based OCT studies on IRDs: Camino et al. [15] investigated the deep learning-based segmentation of preserved photoreceptors on OCT images in the two IRD subtypes choroideremia and retinitis pigmentosa. Zhao et al. [16] developed a few shot-learning approaches to classify OCT images of IRDs into disease subtypes. Miere et al. [17] used a convolutional neural network (CNN) to classify IRDs into subtypes by analyzing fundus autofluorescence images.

Deep Learning algorithms usually require a huge training set, or at least a sufficient number of train images, to result in good generalizable segmentations. However, in our study we exclusively incorporated meticulously chosen OCT volumes, which were characterized by their high quality in terms of having precise annotations. Therefore, we reason that our model performs very well on unseen data, as it was trained on two high quality datasets (healthy and IRD). This achievement was made possible due to the close collaboration between clinicians and AI experts. Subsequently, we could rely on a model [9] which has shown to work well without resorting to more complex architectures or deeper networks, clearly highlighting the strength of our results.

OCT imaging has led to an abrupt and intense advancement of ophthalmological research, as it enabled retinal in-vivo imaging in a resolution that was formerly only known from histological tissue microscopy. While structural abnormalities such as intraretinal fluid accumulations are monitored in exudative retinal diseases, in IRDs, the focus lies on monitoring slowly progressive tissue loss which can be followed up over years. In IRDs of the retinitis pigmentosa type, the outer retinal layers are affected while the inner retinal layers are typically preserved. For IRDs, the segmentation of the outer retinal layers is essential to monitor structural degeneration over time and has been also applied in animal models of preclinical studies [18]. The preservation of outer retinal layers, especially the outer nuclear layer (ONL), in OCT scans has been shown to correlate with residual vision in dark-adapted visual fields in AMD patients [19]. In a study on patients with RPE65-related IRD by Jacobson and colleagues, 96% of retinal loci with residual light sensitivity had a measurable ONL while in 75% of retinal loci without residual sensitivity, no ONL was measurable [20]. Not only the “OCTExplorer’’ used for this study, but also most other commercially available automated segmentation tools fail to reliably segment the distinct layers in OCT scans of patients with IRDs, which results in time consuming manual correction and/or segmentation [19,21].

In 2017, voretigene neparvovec-rzyl (Luxturna), an adeno-associated virus (AAV) vector for gene augmentation therapy in patients with RPE65-related IRD, reached its primary endpoint in a phase III trial which marked a breakthrough in the field of gene therapy [22]. However, since then, other phase III trials on ocular gene therapy have failed to reach their primary endpoints, e.g., the XIRIUS study of an adeno-associated virus serotype 8 (AAV8) vector-based gene therapy, cotoretigene toliparvovec, targeting the RPGR gene in x-linked retinitis pigmentosa. One of the recurring key questions in all ophthalmological gene therapy studies therefore is: “How to measure success?” [23].

This question underlines the complexity and importance of a reliable and objective measurement of visual performance for gene therapy studies in IRDs and beyond. A huge advantage in the diagnostic toolbox for IRDs would be a correlation of structural OCT measurements and functional data. For structure-function prediction, reliable retinal layer segmentation is regarded as crucial. Hence segmentation algorithms like the one presented in this manuscript could play an important role in future studies.

There are limitations of this study. In contrast to AMD or diabetic retinopathy, IRDs are rare diseases, which limits the sample size. Training of the AI was based on manual segmentation of image stacks by experienced ophthalmologists. Manual segmentation bears the risk of human error and may differ slightly between graders. To manually segment the OCT scans for AI training datasets, a minimum resolution quality is needed.

However, in real life settings, imaging resolution in IRD patients may be low due to fixation problems and blinking artifacts. The present study was only performed with Heidelberg Spectralis OCT scans, further investigations should confirm this AI-based tool with scans of other commercially available devices.

In conclusion, AI-based tools are thought to be the key to boost efficiency for an objective assessment of rare diseases, where well-powered randomized controlled trials are almost impossible to conduct due to the limited sample size. The here-presented segmentation and thickness map tool may be a first step to AI-supported IRD diagnostics. It could enable structural disease monitoring in future gene therapy trials and provide the basis for predictive structure-functional modelling.

## Conclusion (bullet points)

### Already known

- Artificial intelligence has an increasingly important role in ophthalmology. To date, most research has focused on high prevalence ophthalmic diseases.
- Our study addresses a critical relevant problem, focusing on reliable retinal layer segmentation for IRD patients. Accurate segmentation of anatomical layers in OCT scans plays a key role in the correlation of retinal structure to visual function.

### Newly described

- We have developed a deep learning algorithm that allows accurate segmentation of pathologically altered OCT scans in patients with inherited retinal diseases and generates a retinal thickness map.
- Future work will explore calculating retinal layer thickness and correlating it with functional data like visual fields. Our aim is to contribute to a greater understanding of disease, and to improve the evaluation of treatment outcomes in future.

## Data Availability

All data produced in the present study are available upon reasonable request to the authors.

## Data Availability

All data produced in the present study are available upon reasonable request to the authors.

## References

[1] Aleman TS, Cideciyan A V., Sumaroka A, et al. Retinal laminar architecture in human retinitis pigmentosa caused by Rhodopsin gene mutations. Invest Ophthalmol Vis Sci 2008; 49: 1580–1591. doi:10.1167/IOVS.07-1110

[2] Apushkin MA, Fishman GA, Alexander KR, et al. Retinal thickness and visual thresholds measured in patients with retinitis pigmentosa. Retina 2007; 27: 349–357. doi:10.1097/01.IAE.0000224944.33863.18

[3] Abràmoff MD, Lavin PT, Birch M, et al. Pivotal trial of an autonomous AI-based diagnostic system for detection of diabetic retinopathy in primary care offices. npj Digit Med 2018 11 2018; 1: 1–8. doi:10.1038/s41746-018-0040-6

[4] Lee CS, Baughman DM, Lee AY. Deep Learning Is Effective for Classifying Normal versus Age-Related Macular Degeneration OCT Images. Ophthalmol Retin 2017; 1: 322–327. doi:10.1016/J.ORET.2016.12.009

[5] Li Z, He Y, Keel S, et al. Efficacy of a Deep Learning System for Detecting Glaucomatous Optic Neuropathy Based on Color Fundus Photographs. Ophthalmology 2018; 125: 1199–1206. doi:10.1016/J.OPHTHA.2018.01.023

[6] Abramoff MD, Garvin MK, Sonka M. Retinal imaging and image analysis. IEEE Rev Biomed Eng 2010; 3: 169–208. doi:10.1109/RBME.2010.2084567

[7] Li K, Wu X, Chen DZ, et al. Optimal Surface Segmentation in Volumetric Images— A Graph-Theoretic Approach. IEEE Trans Pattern Anal Mach Intell 2006; 28: 119. doi:10.1109/TPAMI.2006.19

[8] Garvin MK, Abràmoff MD, Wu X, et al. Automated 3-D intraretinal layer segmentation of macular spectral-domain optical coherence tomography images. IEEE Trans Med Imaging 2009; 28: 1436–1447. doi:10.1109/TMI.2009.2016958

[9] Yu Z, Han X, Xu W, et al. A generalizable brain extraction net (BEN) for multimodal MRI data from rodents, nonhuman primates, and humans. Elife 2022; 11. doi:10.7554/ELIFE.81217

[10] Ting DSW, Pasquale LR, Peng L, et al. Artificial intelligence and deep learning in ophthalmology. Br J Ophthalmol 2019; 103: 167–175. doi:10.1136/BJOPHTHALMOL-2018-313173

[11] Moraes G, Fu DJ, Wilson M, et al. Quantitative Analysis of OCT for Neovascular Age-Related Macular Degeneration Using Deep Learning. Ophthalmology 2021; 128: 693–705. doi:10.1016/J.OPHTHA.2020.09.025

[12] Schmidt-Erfurth U, Vogl WD, Jampol LM, et al. Application of Automated Quantification of Fluid Volumes to Anti–VEGF Therapy of Neovascular Age-Related Macular Degeneration. Ophthalmology 2020; 127: 1211–1219. doi:10.1016/j.ophtha.2020.03.010

[13] Hogarty DT, Mackey DA, Hewitt AW. Current state and future prospects of artificial intelligence in ophthalmology: a review. Clin Experiment Ophthalmol 2019; 47: 128–139. doi:10.1111/CEO.13381

[14] Balyen L, Peto T. Promising artificial intelligence–machine learning–deep learning algorithms in ophthalmology. Asia-Pacific J Ophthalmol 2019; 8: 264–272. doi:10.22608/APO.2018479

[15] Camino A, Wang Z, Wang J, et al. Deep learning for the segmentation of preserved photoreceptors on en face optical coherence tomography in two inherited retinal diseases. Biomed Opt Express 2018; 9: 3092. doi:10.1364/BOE.9.003092

[16] Qi ZHAO, Si Wei MAI, Qian LI, et al. Automated Classification of Inherited Retinal Diseases in Optical Coherence Tomography Images Using Few-shot Learning. Biomed Environ Sci 2023; 36: 431–440. doi:10.3967/BES2023.052

[17] Miere A, Le Meur T, Bitton K, et al. Deep Learning-Based Classification of Inherited Retinal Diseases Using Fundus Autofluorescence. J Clin Med 2020; 9: 1–13. doi:10.3390/JCM9103303

[18] Gardiner KL, Cideciyan A V., Swider M, et al. Long-Term Structural Outcomes of Late-Stage RPE65 Gene Therapy. Mol Ther 2020; 28: 266–278. doi:10.1016/J.YMTHE.2019.08.013

[19] Kelbsch C, Stingl K, Kempf M, et al. Objective Measurement of Local Rod and Cone Function Using Gaze-Controlled Chromatic Pupil Campimetry in Healthy Subjects. Transl Vis Sci Technol 2019; 8: 19–19. doi:10.1167/TVST.8.6.19

[20] Jacobson SG, Aleman TS, Cideciyan A V., et al. Defining the residual vision in leber congenital amaurosis caused by RPE65 mutations. Invest Ophthalmol Vis Sci 2009; 50: 2368–2375. doi:10.1167/IOVS.08-2696

[21] Gersch J, Hufendiek K, Delarocque J, et al. Investigation of Structural Alterations in Inherited Retinal Diseases: A Quantitative SD-OCT-Analysis of Retinal Layer Thicknesses in Light of Underlying Genetic Mutations. Int J Mol Sci 2022; 23: 16007. doi:10.3390/IJMS232416007/S1

[22] Russell S, Bennett J, Wellman JA, et al. Efficacy and safety of voretigene neparvovec (AAV2-hRPE65v2) in patients with RPE65-mediated inherited retinal dystrophy: a randomised, controlled, open-label, phase 3 trial. Lancet (London, England) 2017; 390: 849. doi:10.1016/S0140-6736(17)31868-8

[23] Stingl K, Kempf M, Jung R, et al. Therapy with voretigene neparvovec. How to measure success? Prog Retin Eye Res 2023; 92. doi:10.1016/J.PRETEYERES.2022.101115

